# Cardiovascular, respiratory and functional effects of tele-supervised home-based exercise training in individuals recovering from COVID-19 hospitalization: A randomized clinical trial

**DOI:** 10.1101/2022.01.24.22269745

**Authors:** Vanessa Teixeira do Amaral, Ariane Aparecida Viana, Alessandro Domingues Heubel, Stephanie Nogueira Linares, Bruno Martinelli, Pedro Henrique Camprigher Witzler, Gustavo Yudi Orikassa de Oliveira, Gabriel de Souza Zanini, Audrey Borghi Silva, Renata Gonçalves Mendes, Emmanuel Gomes Ciolac

## Abstract

Our aim was to test the hypothesis that tele-supervised home-based exercise training (exercise) is an effective strategy for improving cardiovascular, respiratory, and functional capacity parameters in individuals that were hospitalized due to coronavirus disease 2019 (COVID-19). Thirty-two individuals (52 ± 10 years; 17F) randomly assigned to exercise (*N* = 12) and control groups (*N* = 20), had their anthropometric (weight, body mass index), hemodynamic (brachial and central blood pressure), vascular (arterial stiffness), ventilatory (pulmonary function and respiratory muscle strength), and functional parameters (handgrip strength, five-time sit to stand [FTSTS], timed up and go test [TUG] and six-minute walking test [6MWT]) assessed at baseline (30 to 45 days of hospital discharged) and after 12 weeks of follow-up. Both groups similarly increased (*P* < 0.001) forced vital capacity (absolute and % of predicted), forced expiratory volume in the first second (absolute and % of predicted), and handgrip strength during follow-up. However, only exercise group reduced carotid-femoral pulse wave velocity (−2.0 ± 0.6 m/s, *P* = 0.048), and increased (*P* < 0.05) resting oxygen saturation (1.9 ± 0.6 %), mean inspiratory pressure (24.7 ± 7.1 cmH_2_O), mean expiratory pressure (20.3 ± 5.8 cmH_2_O) and % of predicted mean expiratory pressure (14 ± 22 %) during follow-up. No significant changes were found in any other variable during follow-up. Present findings suggest that tele-supervised home-based exercise training can a potential adjunct therapeutic to rehabilitate individuals that were hospitalized due to COVID-19.

## 1. INTRODUCTION

The pandemic of coronavirus disease 2019 (COVID-19) is an unprecedented public health emergency, with the exponential increase of cases overloading the health systems worldwide ^1^. Although most COVID-19 cases are mild or even asymptomatic, nearly 20% of patients require hospitalization due to severe manifestations ^2^. The long-term effects of COVID-19 on respiratory, cardiovascular and functional systems is not completely known. However, even under adequate medical treatment, the pneumonia caused by severe acute respiratory syndrome coronavirus 2 (SARS-CoV-2) may cause an injury to the lung parenchyma, with permanent structural damage ^3^. Studies have also shown a direct relationship between COVID-19 and poor cardiovascular outcomes, such as increased arterial stiffness and association between overweight and endothelial dysfunction ^4,5^. Initial investigations showed that SARS-CoV-2 is able to infect the endothelial cells, which are responsible for regulating vascular tone ^6^ harming the vascular function of these individuals ^7^.

To avoid further damage in the long term, interventions aiming to rehabilitate and/or promote the health of COVID-19 patients after hospital discharge are welcome. Exercise training is a well-known first-line intervention for preventing and treating different diseases ^8,9^. Tele-supervised home-based exercise programs have been recommended for promoting health and rehabilitation in different conditions during COVID-19 pandemic ^10^, and may also be a suitable strategy for rehabilitating COVID-19 patients ^10^. However, randomized clinical trials investigating the benefits of tele-supervised home-based exercise training in individuals that were hospitalized due to COVID-19 are still lacking.

Thus, our aim was to test the hypothesis that tele-supervised home-based exercise training is an effective strategy for improving cardiovascular, respiratory, and functional capacity parameters in individuals that were hospitalized due to COVID-19.

## 2. METHODS

### 2.1. Study Design and population

This is a randomized, single center and single-blinded clinical trial (Brazilian Register of Clinical Trials identifier: RBR-9y32yy) that analyzed the effect of a 12-week tele-supervised home-based exercise training on anthropometric, respiratory, cardiovascular and functional parameters in individuals hospitalized due COVID-19. We investigated patients of both sex that were hospitalized at the Bauru State Hospital (São Paulo, Brazil), with age ≥ 18 years, and with laboratory-confirmed COVID-19 diagnosis detected by reverse transcriptase-polymerase chain reaction (RT-PCR) test. Pregnant or lactating women, individuals with contraindications for physical activity (i.e., recent myocardial infarction, unstable angina or arrhythmias or other uncontrolled heart disease), and individuals with decompensated metabolic, pulmonary, hepatic or renal diseases were not included. All volunteers who met inclusion criteria were randomly assigned to perform a 12-week tele-supervised home-based exercise training (exercise) or control follow. The clinical status, anthropometric (body mass, height and body mass index [BMI]), hemodynamic (heart rate [HR], resting systolic and diastolic blood pressure [BP]), vascular (pulse wave velocity [PWV]), ventilatory parameters (pulmonary function and respiratory muscle strength) and physical and functional capacities (handgrip strength, five-time sit to stand [FTSTS], timed up and go test [TUG] and six-minute walking test [6MWT]) were assessed at 30 to 45 days of hospital discharged (baseline) and after 12 weeks of exercise or control follow-up.

Sixty-three individuals who had been hospitalized (in ward setting) due COVID-19 from July 2020 to February 2021, accepted to participate in the study. Two individuals were not included due to decompensated comorbidities. Six-one individuals were then randomly assigned to exercise (*N* = 32) or control (*N* = 29) groups; however, 27 individuals were lost to follow-up due to different reasons. Thus, 32 individuals (exercise = 12 individuals / control = 20 individuals) underwent baseline and follow-up assessments and were included in final analysis (Figure 1). The Ethics Committee of the São Paulo State University (School of Sciences) approved all procedure (CAAE: 32134720.4.1001.5398) and all volunteers provided written informed consent.

**Figure 1:**
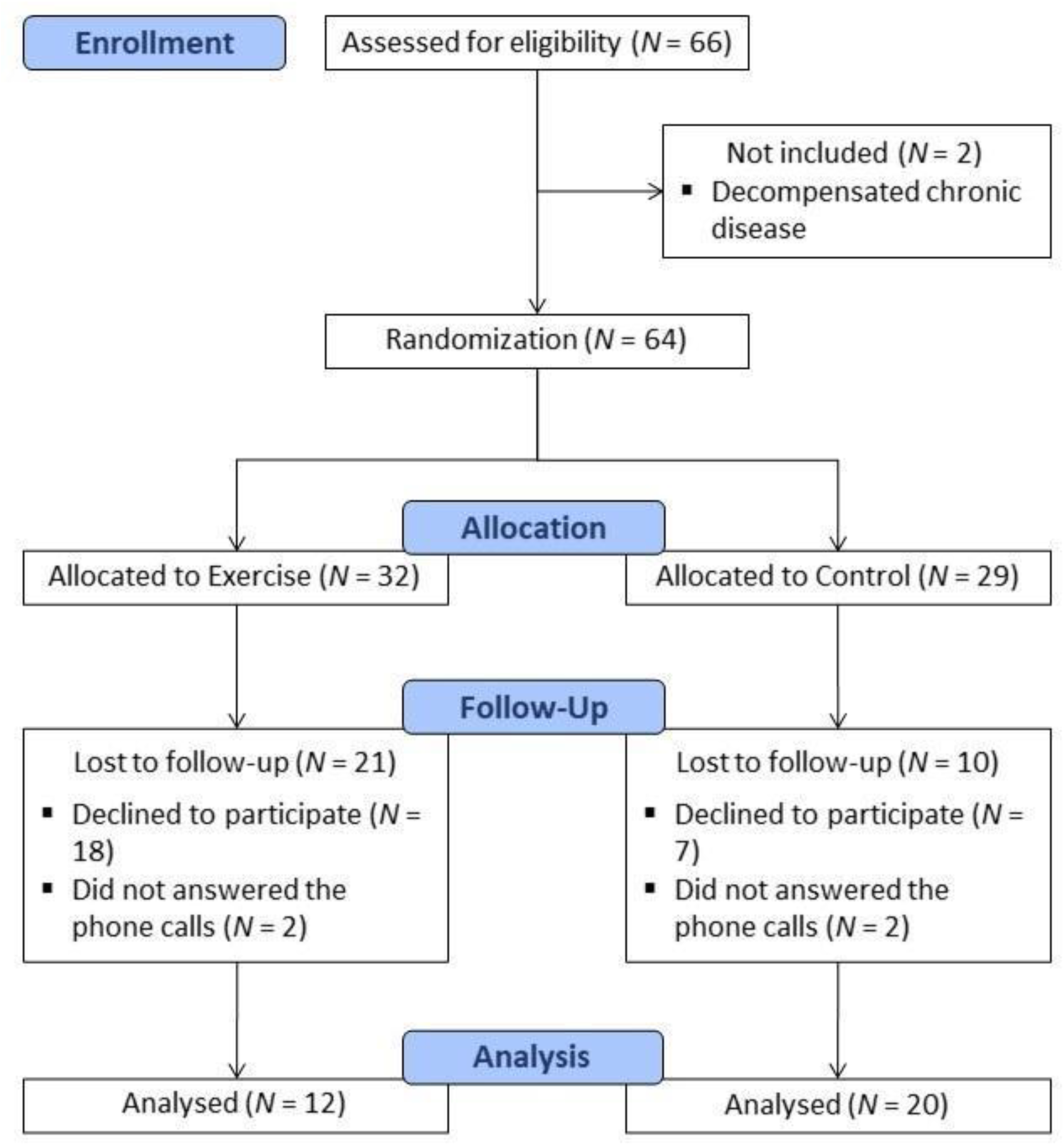
Flowchart of study design.

### 2.2. Clinical assessment

All individuals were invited, by telephone call, to attend Exercise Chronic Disease Laboratory (ECDR) at UNESP/Bauru (SP) to perform both baseline (30 to 45 days after hospital discharge) baseline and follow-up clinical assessments. All measurements were performed in a controlled room temperature (20-22ºC) by the same and experienced evaluator, who was blinded to participant’s group assignment. Clinical assessment included anamnesis (to obtain demographic and health characteristics, persistent symptoms and medications used), body mass and height (Ramuza™ anthropometric scale; Ramuza *Indústria e Comércio de Balanças* Ltda., Santana do Parnaíba-SP, Brazil), and vital signs, respectively. Current smokers were defined as patients who were smoking at the time of study or had stopped smoking during the last month prior its beginning. BMI was calculated using the formula body mass/height^2^ (kg/m^2^) ^13^. Vital signs measurements were performed at seated position, after 10 min of rest, and included pulse oxygen saturation (SpO_2_) (G-Tech™ Led finger oximeter; Accumed *Produtos Médico Hospitalares* Ltda., Duque de Caxias-RJ, Brazil), respiratory rate, BP (Omron HEM 7200™, Omron Healthcare Inc., Dalian, China) and HR (Polar™ H10 heart rate sensor; Polar Electro Inc, Kempele, Finland). SpO_2_ and respiratory were measured once, and BP and HR were measured in triplicated (the average was considered the resting BP and HR), as previously described ^14^.

### 2.3. Arterial stiffness and central pressure

Arterial stiffness and central pressure were assessed at baseline and during follow-up, after clinical assessment, and using a non-invasive automatic equipment (Complior Analyse™ PWV and Central Pressure Analysis ™; Alam Medical, Saint-Quentin-Fallavier, France). The participants were positioned in supine position, and common carotid and femoral arteries pressure waveforms were recorded noninvasively using a pressure-sensitive transducer. The distance between the recording sites (D) was measured in a straight line with a flexible meter, and inserted in the equipment’s software before waveforms measurements. PWV (calculated as PWV = D/t, where (t) means pulse transit time), augmentation index (AIx, ratio of augmentation pressure expressed as the difference between the second and first pressure peaks in the pulse wave) and central pressure (assessed directly from the carotid pressure waveform, using mean and diastolic pressures to calibrate the carotid signal) were automatically calculated ^15^. Pressure waveforms were measured during 10 to 15 cardiac cycles, and the mean was used for the final analysis ^15^. All measurements were performed by an experienced observer that was blinded to participants’ group assignment.

### 2.4. Pulmonary function and respiratory muscle strength

The pulmonary function testing (spirometry) was performed without bronchodilator, using a calibrated and validated portable spirometer (SpiroPro®, Jaeger, Höchberg, Germany), and with participants seated at rest. Forced vital capacity (FVC), forced expiratory volume in the first second (FEV_1_) and peak expiratory flow were obtained by asking the individual for inspiration until total lung capacity and a quick and intense expiration for at least 6 seconds. At least three trials were performed and the largest values of FVC and FEV_1_ were determined. All maneuvers were checked for acceptability and reproducibility criteria ^16^. FVC and FEV_1_ were adjusted to predicted values (FVC % pred and FEV_1_ % pred) according to the Brazilian Guidelines for Pulmonary Function Testing ^17^.

Respiratory muscle strength was measured by analog manovacuometer (Commercial Médica™, São Paulo-SP, Brazil), after pulmonary function testing, and with participants seated at rest. The maximal inspiratory pressure (MIP) was measured with a scale of ± 120 cmH_2_O from residual volume up to the total lung capacity. The maximal expiratory pressure (MEP) was assessed from the total lung capacity, with the individual being instructed to fully inhale and exhale with maximum effort. At least three consecutive trials were carried out, with an interval of one minute between them. The value considered was the highest among the three measurements (except if it was the last), and the predicted values (MIP % pred and MEP % pred) were also calculated ^18^.

### 2.5. Functional capacity

Handgrip strength was measured using a hydraulic dynamometer (Jamar™ hydraulic hand dynamometer, Sammons Preston, Bolingbrook, Illinois, USA) with the individuals in the sitting position, with the elbow flexed at 90º and a neutral wrist. Three measurements were made for the dominant hand. The mean was calculated and the highest value was used for analysis ^19^. Lower limb muscle strength/power was measured by the FTSTS test, after handgrip strength assessment, as previously described ^20^. Balance/agility was measured by the TUG test, after the FTSTS test, as previously described ^20^.

Finally, the 6MWT was assessed on a 30 m length flat surface, using cones and tape measure to mark the ground, and following the recommendations of the European Respiratory Society/American Thoracic Society ^21^. BP was measured before, immediately after and after 2 min of recovery. HR and SpO_2_ were measured before, every 2 min of exercise (2, 4 and 6 min), and at 1 min of recovery. The average of HR and SpO_2_ measured every 2 min of exercise were considered exercise heart rate and exercise SpO_2_, respectively. Absolute (total distance walked during test) and relative (percentage of predicted distance) ^22^ values were used to assess walking performance. The prevalence of partial oxygen desaturation during the exercise phase was measured as a reduction ≤ 4 % in SpO_2_ during any moment of walking when compared to pre-exercise levels.

### 2.6. Exercise training protocol

Participants of the exercise group underwent a 12-week (tele-supervised and home-based) exercise training protocol. At the end of baseline evaluations, exercise group participants received instructions by a trained researcher (an exercise specialist) on how to safely perform the recommended exercise at home. The researcher demonstrated and oriented the participants on how to execute each proposed exercise properly. During this session, the participants were familiarized with each exercise and with the 6 to 20 rating of perceived exertion scale (RPE), which was used to control the exercise intensity ^23^. If necessary, exercise adaptations to properly execute the exercise and/or to meet intensity were made during this session. Supplementary material containing exercise cards, with illustrations and instruction on how to perform each exercise adequately, how to meet the adequate workload (number of repetitions/sets, duration, rhythm, rest interval…) and how to progress workload throughout the follow-up, were sent by mobile app (WhatsApp) immediately after the instructional session. An instructional video about how to properly perform each exercise was also made available on the YouTube platform, and the participants were instructed to watch it as many times as necessary. All participants were contacted individually every Friday (by phone call or WhatsApp messages, according to participants’ preference) to check the exercise frequency, which was noted in a spreadsheet. Verbal encouragements and orientations (if needed) were also performed during the weekly contact.

All participants were instructed to perform both resistance (thrice-weekly in alternated days) and aerobic (five times-a-week) exercises. Participants were also instructed to perform a 5 min warm-up (joint mobility and stretching exercises) and 5 min of cool-down (stretching and relaxing exercises) before and after each exercise session (both resistance and/or aerobics). Resistance training included nine multi-and single-joint exercises (bodyweight squat, push-up on the wall, bodyweight lunge, one-arm row, deadlift, side lateral raise or shoulder press, elbow flexion, calf raise, and abdominal crunch in chair) using bodyweight and/or rubber bands/plastic bottles (with water or sand) as resistance. Participants were instructed to perform 1 set of 10-15 reps at week 1, 2 sets of 10-15 reps at weeks 2 to 3, 3 sets of 10-15 reps at weeks 4 to 6), and 3 sets of 15-20 reps at weeks 7 to 12. Participants were also instructed to maintain 1 min of rest between sets and exercises, and to maintain the intensity at 14-17 points of RPE scale throughout the follow-up.

For aerobic training, participants were instructed to perform walking and/or cycling (depending on preference and equipment available) five times a week, and they may choose to perform part of the sessions after the resistance training or to perform all the sessions in separate days. Aerobic sessions consisted of 10-15 min at week 1 (depending on participant’s capacity), 20 min at weeks 3 to 4, and 30 min at weeks 5 to 12. Exercise intensity should be maintained at 11 to 13 of RPE throughout the follow-up. During weeks 3 to 12, participants could choose to perform the aerobic exercise in a single session (20 or 30 min in a single session) or to accumulate in multiple sessions throughout the day (i.e.; 2 sessions of 10 min at weeks 3 to 4, and 2 sessions of 15 min or 3 sessions of 10 min at weeks 5 to 12).

### 2.7. Statistical analysis

Statistical analysis was performed using the Statistical Package for the Social Sciences version 19.0 (SPSS Inc., Chicago, IL, USA) for Windows. The Shapiro-Wilk and Levene tests were used to assess normality and homoscedasticity of data, respectively. Data were expressed as mean ± SD (parametric data) or as *N* (%) (categorical data). Unpaired Student’s t test and Chi-square were used to indicate difference between groups in parametric and categorical variables at baseline. Two-way analysis of variance (ANOVA) with repeated measures (group vs. time) was used to indicate between-and within-group differences in the variables measured before and during follow-up. The Bonferroni post-hoc test was used to identify the significant differences indicated by ANOVA. The level of significance was set at *P* < 0.05.

## 3. RESULTS

Characteristics of the participants included in the study were not different between groups at baseline (Table 1). Seventy-five percent and 85% of exercise and control group had at least one comorbidity, respectively, and the most prevalent were obesity and hypertension. The tele-supervised home-based exercise program was well tolerated by all participants and there were no adverse events during follow-up. The prevalence of subjects with at least one persistent COVID-19 related symptom was not different between groups both at baseline (exercise = 83%; control = 85%; P = 0.900) and during follow-up (exercise = 75%; control = 85%, *P* = 0.483). However, the quantity of persistent symptoms tended to reduce in exercise (baseline = 2.5 ± 2.4 symptoms; follow-up = 1.6 ± 1.3 symptoms; *P* = 0.085), but not in control (baseline = 2.6 ± 1.5; follow-up = 2.3 ± 1.9; *P* = 0.464). Two-way ANOVA indicated significant within-group difference in body weight (*F*_1, 30_ = 10.237; *P* = 0.003; η^2^ = 0.254; power = 0.872). *Post hoc* analysis identified that body weight increased significantly (2.76 ± 0.62 kg) in control (baseline = 89.0 ± 21.7 kg; follow-up = 90.5 ± 22.0 kg; *P* = 0.010), whereas a tendency toward increase (1.87 ± 0.54 kg) occurred in exercise (baseline = 89.1 ± 14.4 kg; follow-up = 90.4 ± 14.1 kg; *P* = 0.063).

**Table 1.** Participant’s characteristics at baseline.

| **Variable** | **Exercise (N = 12)** | **Control (N = 20)** | **P** |
| --- | --- | --- | --- |
| Age (years) | 51.9 ± 10.2 | 53.3 ± 11.6 | 0.506 |
| Sex [female (%)] | 5 (42) | 12 (60) | 0.314 |
| Race (white/black/mixed/indigenous) | 7/4/1/0 | 15/3/1/1 | 0.534 |
| BMI (kg/m<sup>2</sup>) | 32.6 ± 7.4 | 32.4 ± 7.8 | 0.624 |
| Smoking (never/current/former) | 9/1/2 | 15/1/4 | 0.915 |
| Comorbidities [N (%)] | 9 (75) | 17 (85) | 0.647 |
| *Hypertension [N (%)]* | 5 (42) | 11 (55) | 0.716 |
| *Diabetes [N (%)]* | 4 (33) | 1 (5) | 0.053 |
| *Cardiovascular disease [N (%)]* | 0 (0) | 2 (10) | 0.516 |
| *Obesity [N (%)]* | 8 (67) | 13 (65) | 0.923 |
| *Dyslipidemia [N (%)]* | 1 (8) | 2 (10) | 0.876 |
| *Respiratory disease [N (%)]* | 1 (8) | 2 (10) | 0.876 |
| *Hypothyroidism [N (%)]* | 0 (0) | 1 (5) | 1.000 |
| *Others [N (%)]* | 3 (25) | 4 (20) | 0.740 |
| Previous surgeries (No/Yes) | 8 (67) | 16 (80) | 0.399 |
| Hospital stay (days) | 6 ± 2 | 7 ± 5 | 0.117 |
| Adverse event [N (%)] | 0 (0) | 2 (10) | 0.258 |
| *ICU admission [N (%)]* | 0 (0) | 2 (10) | 0.258 |
| *IMV [N (%)]* | 0 (0) | 0 (0) | 1.000 |
| *Cardiovascular [N (%)]* | 0 (0) | 1 (5) | 0.431 |
| *Non-cardiovascular [N (%)]* | 0 (0) | 1 (5) | 0.431 |
| Days post hospital discharge (days) | 35.6 ± 6.6 | 35.7 ± 5.1 | 0.442 |

BMI: body mass index; ICU: intensive care unit; IMV: invasive mechanical ventilation.

Two-way ANOVA also indicated within and between group interactions in resting SpO_2_ (*F*_1, 30_ = 5.455; *P* = 0.026; η^2^ = 0.154; power = 0.618), and within group difference in PWV (*F*_1, 28_ = 6.129; *P* = 0.020; η^2^ = 0.180; power = 0.666), but no significant differences were indicated in any other hemodynamic and vascular variables (Table 2). *Post hoc* analysis identified that exercise group, but not control group, increased resting SpO_2_ (1.9 ± 0.6; *P* = 0.015) (Table 2) and reduced PWV (−2.0 ± 0.6 m/s; *P* = 0.043) (Figure 2) during follow-up.

**Table 2.** Resting respiratory, hemodynamic, and vascular variables at baseline and during follow-up.

| Variable | Exercise (N = 12) |  | Control (N = 20) |  |
| --- | --- | --- | --- | --- |
|  | Baseline | Follow-up | Baseline | Follow-up |
| Respiratory rate (rpm) | 14 ± 4 | 15 ± 3 | 16 ± 5 | 18 ± 3 |
| SpO<sub>2</sub> (%) | 96.0 ± 1.0 | 97.0 ± 1.6 * | 96.7 ± 1.1 | 96.6 ± 1.1 |
| Heart rate (bpm) | 83 ± 13 | 80 ± 13 | 76 ± 14 | 77 ± 13 |
| Brachial blood pressure |  |  |  |  |
| *Systolic (mmHg)* | 129 ± 24 | 126 ± 16 | 126 ± 22 | 125 ± 20 |
| *Diastolic (mmHg)* | 83 ± 10 | 82 ± 8 | 77 ± 13 | 75 ± 9 |
| Augmentation index | 11.5 ± 18.1 | 8.2 ± 13.9 | 15.7 ± 14.5 | 8.6 ± 17.2 |
| Central blood pressure |  |  |  |  |
| *Systolic (mmHg)* | 121 ± 22 | 118 ± 21 | 115 ± 20 | 114 ± 18 |
| *Diastolic (mmHg)* | 84 ± 11 | 82 ± 9 | 77 ± 13 | 75 ± 10 |
| *Pulse (mmHg)* | 38 ± 14 | 34 ± 11 | 38 ± 14 | 40 ± 13 |
| *Pulse pressure ratio* | 1.26 ± 0.26 | 1.29 ± 0.21 | 1.32 ± 0.19 | 1.27 ± 0.19 |
| *Augmentation pressure* | 7.4 ± 6.6 | 4.2 ± 3.7 | 6.0 ± 5.2 | 6.1 ± 4.4 |

Data are presented as mean ± SD. SpO<sub>2</sub>: oxygen saturation. Asterisk denotes significant difference from baseline at the same group (\*: $P < 0.05$ ).

**Figure 2.**
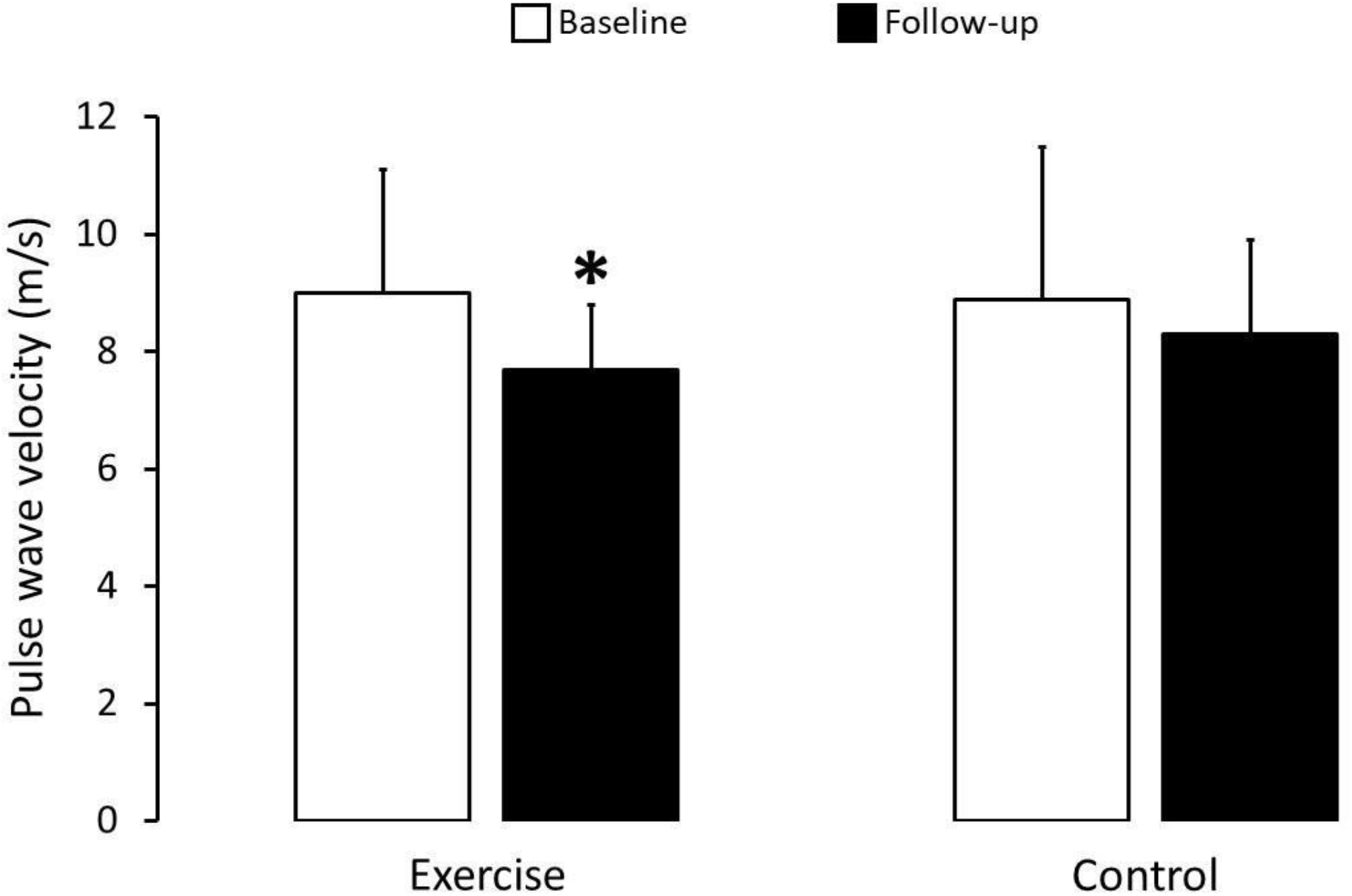
Pulse wave velocity at baseline and during follow-up. Asterisk denotes significant difference from baseline at the same group (*: *P* = 0.043).

In addition, two-way ANOVA indicated within group differences in FVC (*F*_1, 30_ = 33.727; *P* < 0.001; η^2^ = 0.529; power = 1.0), FVC % pred (*F*_1, 30_ = 36.111; *P* < 0.001; η^2^ = 0.546; power = 1.0), FEV_1_ (*F*_1, 30_ = 28.673; *P* < 0.001; η^2^ = 0.489; power = 0.999), FEV_1_ % pred (*F*_1, 30_ = 31.416; *P* < 0.001; η^2^ = 0.512; power = 1.0), peak expiratory flow (*F*_1, 30_ = 5.119; *P* < 0.031; η^2^ = 0.146; power = 0.591), MIP (*F*_1, 30_ = 22.065; *P* < 0.001; η^2^ = 0.441; power = 0.995), MIP % pred (*F*_1, 28_ = 18.380; *P* < 0.001; η^2^ = 0.396; power = 0.985), MEP (*F*_1, 28_ = 21.810; *P* < 0.001; η^2^ = 0.438; power = 0.995), and MEP % pred (*F*_1, 28_ = 17.684; *P* < 0.001; η^2^ = 0.387; power = 0.982). ANOVA did not indicate significant differences in FEV_1_ / FVC ration. *Post hoc* analysis identified that both groups increased (*P* < 0.001) FVC (exercise: 0.43 ± 0.12 l; control: 0.48 ± 0.11 l), FVC % pred (exercise: 14 ± 12 %; control: 12 ± 12 %; ∼ 0.5 and ∼ 0.5), FEV_1_ (exercise: 0.46 ± 0.13 l; control: 0.36 ± 0.08 l), and FEV_1_ % pred (exercise: 15 ± 15 %; control: 11 ± 12 %; ∼ 0.5 l and ∼ 0.3 l) during follow-up (Figure 3). However, only exercise group increased MIP (24.7 ± 7.1 cmH_2_O, *P* < 0.001), MEP (20.3 ± 5.8 cmH_2_O, *P* = 0.021), and MEP % pred (14.3 ± 22.6 %, *P* = 0.042) during follow-up (Figure 3). No significant differences were identified in peak expiratory flow and MIP % pred.

**Figure 3.**
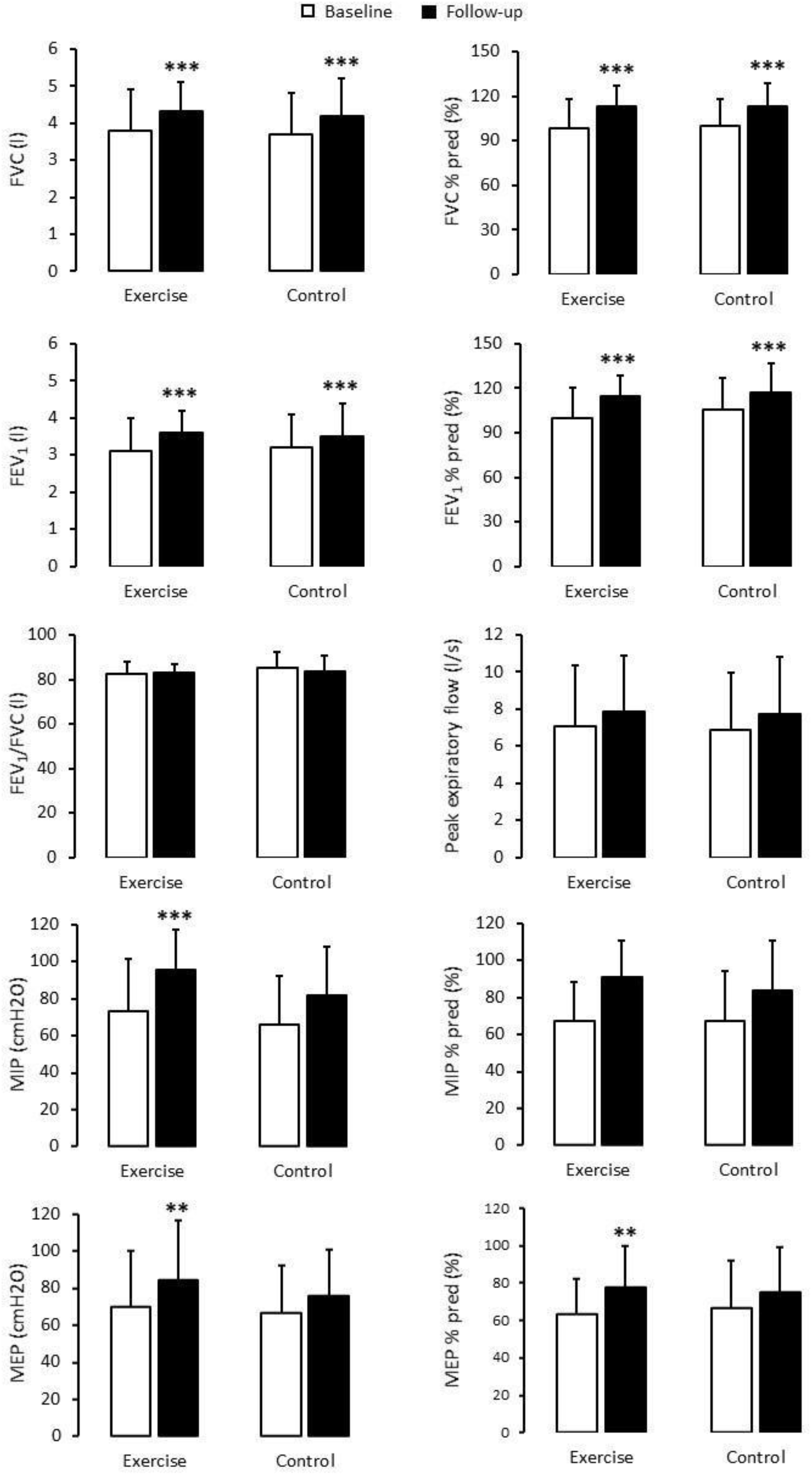
Pulmonary function and respiratory muscle strength at baseline and during follow-up. FVC: forced vital capacity; FVC % pred: percentage of predicted forced vital capacity; FEV_1_: forced expiratory volume in the first second; FEV_1_ % pred: percentage of predicted forced expiratory volume in the first second; MIP: maximal inspiratory pressure; MIP % pred: percentage of predicted maximal inspiratory pressure; MEP: maximal expiratory pressure; MEP % pred: percentage of predicted maximal expiratory pressure. Asterisk denotes significant difference from baseline at the same group (**: *P* < 0.01; ***: *P* < 0.001).

Indeed, two-way ANOVA indicated within group difference in handgrip (*F*_1, 30_ = 341.901; *P* < 0.001; η^2^ = 0.349; power = 0.972) and FTSTS (*F*_1, 30_ = 4.465; *P* < 0.043; η^2^ = 0.130; power = 0.534), as well as SpO_2_ at pre (*F*_1, 29_ = 8.088; *P* = 0.008; η^2^ = 0.218; power = 0.785) and exercise (*F*_1, 29_ = 35.048; *P* < 0.001; η^2^ = 0.547; power = 1.0) phases of 6MWT. ANOVA also indicated significant between group differences in TUG (*F*_(1, 30)_ = 4.817; *P* < 0.036; η^2^ = 0.138; power = 0.565), and HR during exercise (*F*_1, 29_ = 9.415; *P* = 0.005; η^2^ = 0.245; power = 0.843) and recovery (*F*_1, 29_ = 5.001; *P* = 0.033; η^2^ = 0.147; power = 0.580) phases of 6MWT. No significant differences were indicated in any other variable of 6MWT (Table 3). *Post hoc* analysis identified that both groups increased (*P* < 0.05) handgrip strength (exercise: 4.5 ± 1.3 kgf; control: 4.6 ± 1.0 kgf) and SpO_2_ at exercise phase of 6MWT during follow-up, while SpO_2_ at pre 6MWT increased (*P* = 0.018) only in exercise (Table 3). *Post hoc* analysis also identified that HR at exercise and recovery phases of 6MWT were higher (*P* < 0.05) in exercise than control group (both at baseline and during follow-up). No significant differences during follow-up were identified in FTSTS, and no significant difference between groups were identified in TUG (Table 3).

**Table 3.** Functional capacity at baseline and during follow-up.

| Variable | Exercise (N = 12) |  | Control (N = 20) |  |
| --- | --- | --- | --- | --- |
|  | Baseline | Follow-up | Baseline | Follow-up |
| Handgrip (kgf) | 37.5 ± 11.2 | 40.8 ± 12.0 * | 31.2 ± 11.0 | 35.1 ± 10.3 ** |
| FTSTS (s) | 12.7 ± 4.5 | 11.2 ± 2.8 | 13.9 ± 3.5 | 12.3 ± 2.4 |
| TUG (s) | 6.6 ± 1.7 | 6.3 ± 1.4 | 7.6 ± 1.3 | 7.2 ± 1.1 |
| 6MWT distance (m) | 522 ± 107 | 543 ± 115 | 472 ± 98 | 490 ± 76 |
| 6MWT relative distance (% of predicted) | 92.2 ± 14.3 | 96.0 ± 16.4 | 85.7 ± 16.7 | 89.1 ± 13.5 |
| 6MWT Heart rate |  |  |  |  |
| *Pre (bpm)* | 84 ± 12 | 82 ± 10 | 78 ± 14 | 78 ± 14 |
| *Exercise (bpm)* | 118 ± 18 | 118 ± 19 | 100 ± 14 †† | 106 ± 14 † |
| *Recovery (bpm)* | 90 ± 14 | 89 ± 14 | 81 ± 12 † | 79 ± 12 † |
| 6MWT systolic blood pressure |  |  |  |  |
| *Pre (mmHg)* | 129 ± 24 | 126 ± 16 | 123 ± 17 | 125 ± 20 |
| *Exercise (mmHg)* | 151 ± 24 | 158 ± 17 | 147 ± 29 | 151 ± 25 |
| *Recovery (mmHg)* | 134 ± 20 | 135 ± 17 | 137 ± 25 | 134 ± 18 |
| 6MWT diastolic blood pressure |  |  |  |  |
| *Pre (mmHg)* | 83 ± 10 | 82 ± 8 | 76 ± 13 | 74 ± 10 |
| *Exercise (mmHg)* | 90 ± 14 | 93 ± 11 | 87 ± 18 | 88 ± 16 |
| *Recovery (mmHg)* | 90 ± 14 | 93 ± 14 | 87 ± 15 | 88 ± 15 |
| 6MWT SpO<sub>2</sub> |  |  |  |  |
| *Pre (%)* | 96.7 ± 1.1 | 97.6 ± 1.1 ** | 97.2 ± 1.1 | 97.5 ± 1.2 |
| *Exercise (%)* | 95.0 ± 1.5 | 96.4 ± 0.8 ** | 95.2 ± 1.5 | 96.3 ± 1.0 *** |
| *Recovery (%)* | 96 ± 2 | 97 ± 1 | 97 ± 1 | 97 ± 1 |

Data are presented as mean ± SD; FTSTS: five-time sit to stand; SpO<sub>2</sub>: oxygen saturation; TUG: timed up and go test. Asterisk denotes significant difference from baseline at the same group (\*\*: $P < 0.01$ ; \*\*\*: $P < 0.001$ ). Dagger denotes significant difference from exercise at the same moment (†: $P < 0.05$ ; ††: $P < 0.01$ ).

## 4. DISCUSSION

The major finding of the present study was that tele-supervised home-based exercise training was effective to reduce PWV in individuals recovering from COVID-19 hospitalization. In addition, although FVC, FVC % pred, FEV_1_, and FEV_1_ % pred increased similarly in both groups, MIP, MEP, MEP % pred, and resting SpO_2_ only in the exercise group. To our knowledge, this is the first randomized controlled trial to investigate the effect of short-term (12 weeks) exercise training on cardiovascular, respiratory, and functional capacity parameters in individuals recovering from COVID-19 hospitalization.

Current knowledge regarding the underlying pathophysiological mechanisms of vascular dysfunction after SARS-CoV-2 infection is limited ^24^. PWV is a vascular measure that is an important predictor of cardiovascular events and may improve risk reclassification in those at intermediate cardiovascular risk ^25^. For example, a 1 m/s of increase in PWV is associated with a 14 % increased risk in cardiovascular events, and a 15 % increased risk in cardiovascular and all-cause mortality ^26^. Previous studies showed that PWV is increased in individuals infected with SARS-CoV-2 when compared to a control group ^7,27^. The present 2.0 ± 0.6 m/s PWV reduction in exercise group and no significant change in control suggest that tele-supervise home-based exercise training is effective for improving arterial stiffness in survivors from COVID-19 hospitalization, which may impact cardiovascular prognosis and mortality in the long term ^25,26^.

Previous systematic reviews assessing the effect of face-to-face exercise training on arterial stiffness showed an average reduction of only 0.6 m/s in PWV after exercise programs with durations similar to ours (12 weeks) ^28,29^. It can be speculated that the higher PWV decrease found in the present study may be associated with an increased inflammatory process ^30^ caused by COVID-19. Inflammatory process deteriorates vascular integrity, inducing hyperinflammation and release of cytokines ^30^, reducing the bioavailability of nitric oxide and consequently increasing arterial stiffness ^27,30^. This inflammatory process activation is associated with serious damage to target organs, in addition to disruption of endothelial cells ^27^. In accordance, COVID-19 survivors showed increased PWV after four months of infection, which was associated with oxidative stress and endothelial dysfunction markers ^31^. Thus, it is possible that the well-known anti-inflammatory benefits of exercise training ^8^ may revert the COVID-19 inflammatory process, resulting in the present large PWV reduction. Future studies assessing this hypothesis are then welcome.

Evidence about pulmonary function after COVID-19 hospital discharge is currently limited to cross-sectional studies ^32^, where disease severity correlated with reduced pulmonary function 4 months after acute infection ^33^. In the present study, we found similar FVC and FEV_1_ (absolute and relative levels) improvements during follow-up in both exercise and control groups. Considering that these patients had a disease that affects the respiratory system ^32^ and had values within the normal range after hospital discharge, the present finding suggests that the natural process of recovery occurs without the influence of exercise training. Previous studies have also shown reduced levels of respiratory muscle strength (MIP and MEP) after hospital discharge among COVID-19 patients ^33–35^, which is similar to the lower baseline levels of MIP % pred and MEP % pred found in the present study (both exercise and control groups). Thus, individuals hospitalized due to COVID-19 appear to have long-lasting pulmonary parenchymal dysfunction that results in changes in mechanical properties of the chest wall and respiratory muscles.

On the other hand, tele-supervised home-based exercise was effective to improve MIP (24.7 ± 7.1 cmH_2_O), MEP (20.3 ± 5.8 cmH_2_O), MEP % pred (14.3 ± 22.6 %), and resting SpO_2_ (1.9 ± 0.6 %). It is important to note that prevalence of inspiratory or expiratory muscle strength impairment (MIP or MEP levels lower than 80 % of the predicted levels) in exercise group reduced from 8 (67%) and 9 (75%) individuals to 3 (25%) and 5 (42%) for MIP and MEP, respectively. Average levels of MIP and MEP (both absolute and % pred) also increased in the control group during follow-up; however, the increase was of lower magnitude and not statistically significant. Although the present exercise program was not focused on training the respiratory muscles, these muscles are indistinctly activated during exercise, which probably caused the greater respiratory pressures increases in exercise than control group. Accordingly, resistance ^36^ and aerobic ^37^ exercise training programs similar to present study showed to be effective for improving MIP and MEP in other populations. Regarding resting SpO_2_ improvement found only in exercise group, it can be speculated that it is a result of a greater O_2_ delivery and, consequently, availability due to the vascular and respiratory strength improvements induced by the exercise program.

It is also important to note that there was a similar increase in handgrip strength, and no significant changes in FTSTS, TUG and 6MWT distance during follow-up in both groups. In addition, the exercise-induced increase in resting SpO_2_ did not result in improved exercise SpO_2_ when compared to control follow-up. Findings from a previous study assessing the effect of different face-to-face exercise programs (using exercise similar to those used in the present study) in older individuals suggest that a higher aerobic exercise intensity or resistance exercise volume may be required to increase these functional capacity parameters ^38^. Future studies assessing this hypothesis in individuals recovering from COVID-19 are thus welcome.

The small sample size with only non-critical hospitalized individuals does not allow us to extrapolate the present results to all individuals recovering from COVID-19. Although the power and effect sizes were adequate for most of the assessed variables, future studies assessing the effects of exercise in other COVID-19 populations (i.e.; critical and mild disease) are required. The high drop-out rate is also a limitation that should be addressed. When asked by phone, the main reasons for dropping-out included lack of time, to work overtime (to replace co-workers infected by SARS-CoV-2), to take care of family members, and fear of leaving their house and being re-infected by SARS-CoV-2. Future studies and exercise programs addressing to overcome these barriers are thus welcome.

## 5. CONCLUSION

In summary, both groups similarly increased pulmonary function and handgrip strength during follow-up. However, only exercise group reduced carotid-femoral pulse wave velocity, and increased respiratory muscle strength and SpO_2_. These findings suggest that tele-supervised home-based exercise training can be a potential adjunct therapeutic to rehabilitate individuals that were hospitalized due to COVID-19.

## Data Availability

The data used to support the findings of this study are available through the
corresponding author upon reasonable request.

## AVAILABILITY OF DATA

The data used to support the findings of this study are available through the corresponding author upon reasonable request.

## ACKNOWLEDGEMENT

The authors are thankful to Bauru State Hospital (HEB/FAMESP) staff for the assistance with the study. This work was supported by *Fundação de Amparo à Pesquisa do Estado de São Paulo* (FAPESP #2015/09259-2) and *Coordenação de Aperfeiçoamento de Pessoal de Nível Superior* (CAPES – Finance Code 001). Vanessa Teixeira do Amaral was supported by CAPES (Finance Code 001) and EGC was supported by *Conselho Nacional de Desenvolvimento Científico e Tecnológico* (CNPq #303399/2018-0).

## AUTHOR’S CONTRIBUTIONS

VTA, AAV, ADH, BM, RGM and EGC participated in the study conception and design. Material preparation and data collection were performed by VTA, AAV, GYOO, PHCW, ADH, and SNL. Data analysis and interpretation was performed by EGC, and VTA participated in data interpretation. The first manuscript draft was performed by VTA, AAV, GSZ, BM and EGC. All authors contributed to manuscript draft improvement. Final revision was made by VTA and EGC. All authors read and approved the final version of the manuscript.

